# MRI-informed active surveillance outcomes and dietary fibre acceptability: the Aberdeen experience

**DOI:** 10.64898/2026.09.24.26363863

**Authors:** Daniel Sescu, Jakub Zbikowski, Justine S Royle, Sara J MacLennan, Anne E Kiltie

## Abstract

**Objectives:** To describe active surveillance (AS) outcomes following introduction of an MRI-informed pathway, examine whether baseline PI-RADS/Likert scores identify men with earlier AS discontinuation, and assess the acceptability of dietary fibre supplementation in men on AS.

**Patients and methods:** We conducted a retrospective audit of 420 men enrolled in AS at Aberdeen Royal Infirmary between 2012 and 2022. Men were grouped into an MRI-informed pathway (n=198) or biopsy-first pathway (n=222). Time to AS discontinuation for any reason was analysed using Kaplan-Meier methods and log-rank tests. An exploratory analysis compared men with baseline PI-RADS/Likert scores of 4-5 and 2-3. Ten men on AS also completed semi-structured interviews analysed using the Theoretical Framework of Acceptability.

**Results:** Median follow-up was 3.1 years (IQR 1.6-5.1). Overall, 208 men (49.5%) discontinued AS, including 176 because of disease progression. Median time to discontinuation was 4.9 years (95% CI 4.4-5.6). It was shorter in the MRI-informed pathway than in the biopsy-first pathway (4.1 vs 5.6 years; log-rank p<0.01). Among 167 men with recorded baseline MRI scores, 44 (26.3%) had PI-RADS/Likert 4-5. Median time to discontinuation was 2.9 years in this group compared with 4.0 years for scores 2-3 (p<0.01). Interview participants were generally willing to consider dietary fibre supplementation, although gastrointestinal effects and study procedures were anticipated burdens.

**Conclusion:** AS discontinuation occurred earlier in the MRI-informed pathway, although differences in patient selection and surveillance practice should be considered when interpreting this finding. PI-RADS/Likert 4-5 identified a sizeable subgroup with earlier all-cause AS discontinuation that may help inform recruitment to future intervention studies. Dietary fibre supplementation was acceptable in principle, supporting further feasibility work.

## Introduction

Active surveillance (AS) is a standard management option for men with low-risk localised prostate cancer. It can avoid or delay radical prostatectomy or radiotherapy and their associated adverse effects [1,2]. Current UK NICE guidance recommends AS for men with Cambridge Prognostic Group 1 disease and recommends multiparametric magnetic resonance imaging (mpMRI) for men on AS who have not previously undergone MRI [3]. NHS Grampian incorporated mpMRI into its diagnostic and surveillance pathway in 2018, replacing an earlier biopsy-first approach.

Prostate MRI findings are commonly reported using PI-RADS or Likert scores from 1 to 5 [4,5]. Higher scores indicate a greater suspicion of clinically important prostate cancer. In men on AS, the baseline MRI score may help identify those more likely to discontinue surveillance earlier [6]. This is relevant to future dietary intervention studies aimed at delaying progression. Recruiting men more likely to reach an endpoint could reduce the required sample size and trial duration.

Although AS is oncologically safe for appropriately selected men with low-risk localised prostate cancer, some discontinue surveillance because of disease reclassification, patient choice or transition to watchful waiting [2,7]. In a recently published real-world cohort, treatment-free survival decreased with increasing pathological risk. However, most men in that cohort were managed before routine mpMRI, so MRI could not be assessed as a standardised predictor of surveillance outcomes [8]. MRI-visible disease has been associated with earlier AS discontinuation [6]. Further evaluation of baseline PI-RADS/Likert scores may therefore help identify men more likely to discontinue AS earlier and inform recruitment to future intervention studies [6].

Emerging research shows modifiable lifestyle factors, particularly diet, are involved in cancer pathogenesis and progression. The gut microbiota, the community of microorganisms living in the gastrointestinal tract, ferments dietary fibre into short-chain fatty acids (SCFAs), including butyrate. These SCFAs may have anti-tumour effects through mechanisms including histone deacetylase inhibition and altered immune function within the tumour microenvironment [9]. While epidemiological studies consistently highlight the link between higher dietary fibre intake has been associated with reduced colorectal cancer risk [10], while evidence for dietary fibre supplementation in prostate cancer is limited, with studies involving different tumour stages.

In a population-based US study, higher dietary fibre intake was associated with less aggressive prostate cancer [11]. Small low-fat, high-fibre, soy-supplemented dietary intervention studies also reported reduced *ex vivo* LNCaP cell growth following prostatectomy and in untreated localised prostate cancer [12,13]. Keizman et al. reported favourable PSA-related outcomes and good tolerability for modified citrus pectin in men with non-metastatic biochemical recurrence after prior local therapy [14].

More recently, Thomas et al. tested a capsule-based intervention, randomising 208 men with low-risk prostate cancer managed with surveillance to receive phytochemical-rich food capsules with either a five-strain *Lactobacillus* probiotic blend with some inulin fibre or placebo [15]. PSA progression was slower in the probiotic group over four months, with longer follow-up needed to determine whether this leads to clinically meaningful differences in disease progression. However, it is not possible to disentangle the active ingredient(s) within the capsule intervention.

Further research is needed before dietary fibre might be used routinely in prostate cancer care. We do not yet know whether men on AS would find a daily fibre supplement acceptable or practical within a clinical trial.

In this study we aimed to: (1) determine outcomes in the AS population in a single teaching hospital in the north east of Scotland (Aberdeen Royal Infirmary), comparing the modern MRI era with the previous biopsy-first approach, identifying features of men more likely to discontinue AS earlier, and (2) assess the theoretical acceptability for men on AS of taking dietary fibre supplementation in a clinical trial, using structured patient interview-based qualitative research. This study provides information to guide the design of a future dietary fibre intervention study in men on AS to delay time to radical treatment.

## Patients and methods

### Clinical audit

We conducted a single-centre audit of men enrolled in AS at Aberdeen Royal Infirmary, NHS Grampian, between January 2012 and December 2022. Men were eligible if they were aged ≥18 years, had histologically confirmed prostate cancer and had been accepted onto the local AS pathway following multidisciplinary team review. Men who had received definitive treatment before entering AS were excluded. No other exclusion criteria were applied.

The audit was approved by the NHS Grampian Caldicott Guardian for Quality Assurance (reference 15439) and reporting was informed by the STROBE statement [16].

Men were identified from the Urology department database and multidisciplinary team records. Data were extracted from TrakCare EMR (InterSystems) using a standardised proforma. Variables included demographics, prostate-specific antigen (PSA), diagnostic and repeat biopsy findings, MRI findings and reasons for AS discontinuation. A summary of variables collected is provided in the supplementary table S1.

When prostate volume was not reported, it was estimated using the prolate ellipsoid formula:

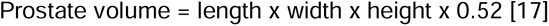

Tumour volume was estimated as the mean percentage involvement across the relevant positive biopsy cores. PSA density was calculated using the PSA value closest to the MRI date and the corresponding prostate volume. PSA doubling time was calculated as In(2) divided by the slope from linear regression of In-transformed PSA against time. At least three measurements from the preceding 12 months were required [18].

The local AS protocol changed during the study period. Under the earlier biopsy-first protocol, diagnosis was based on systematic biopsy without routine pre-biopsy MRI. PSA testing and digital rectal examination were scheduled every 3 months for 2 years, every 6 months to 5 years and annually thereafter. Repeat biopsies were scheduled at years 1,4,7 and 10, then every 5 years. A PSA doubling time of <3 years was a potential reason to discontinue AS. A value of 3-10 years prompted repeat biopsy unless one had been performed within the previous 12 months.

From 2018, pre-biopsy mpMRI was performed whenever possible. PSA was measured every 3 months initially and every 6 months when stable. Clinical reviews were conducted every 6 months. MRI and repeat biopsy were scheduled at 12, 24 and 36 months, with subsequent MRIs and repeat biopsies every two years thereafter. The two pathways are summarised in Supplementary table S2.

Reasons for discontinuation were classified as pathological, biochemical or radiological progression; combined radiological and biochemical progression; patient choice; or transition to watchful waiting. Pathological progression included upgrading to Grade Group ≥2 or increased pathological tumour burden without upgrading. Biochemical progression included increased PSA density or adverse PSA kinetics. Radiological progression included increased lesion size or conspicuity, a new lesion, stage progression or a PRECISE score ≥4 [19].

Men were grouped according to the diagnostic pathway leading to prostate cancer diagnosis. The MRI-informed pathway formally commenced in January 2018 (n=198): 168 underwent pre-biopsy MRI and 30 underwent MRI after a previous negative biopsy but before the biopsy that confirmed cancer. This group included men who started AS from 2018 onwards and two men from late 2017 (despite this pathway formally commencing in January 2018). The biopsy-first pathway included 222 men diagnosed by systematic biopsy without routine pre-biopsy MRI.

### Statistical analysis

Continuous variables were reported as means with standard deviations or medians with interquartile ranges, as appropriate. Categorical variables were reported as frequencies and percentages. Normality was assessed using the Shapiro-Wilk test.

Groups were compared using Student’s *t*-test, Welch’s *t*-test or the Mann-Whitney *U* test for continuous variables, and the chi-squared or Fisher’s exact test for categorical variables.

Kaplan-Meier methods estimated the probability of remaining on AS. Follow-up began at AS enrolment. For time-to-event analyses, the event was AS discontinuation for any reason, using the documented date on which AS ended. Men remaining on AS were censored at their last clinical review, up to 24 April 2024. Deaths before discontinuation were censored at the date of death.

Groups were compared using the log-rank test. All tests were two-sided, and *p*<0.05 was considered statistically significant. Analyses used SPSS version 29 (IBM Corp., Armonk, NY, USA) and Stata Basic Edition version 18.5 (StataCorp LLC, College Station, TX, USA).

Missing values were recorded as unavailable and were not imputed. Men were included in the primary time-to-event analysis if the AS enrolment date, discontinuation status and either a discontinuation or censoring date were available. Missing data were not imputed. The primary time-to-event analysis required an AS enrolment date, AS status and a discontinuation or censoring date. Missing secondary variables affected only analyses requiring those variables.

### Qualitative study

Semi-structured interviews explored the acceptability of dietary fibre supplementation among men on AS. Men were eligible if they were aged _≥_18 years, English-speaking, currently receiving AS and able to use Microsoft Teams. Men were excluded if they had received surgery, radiotherapy, chemotherapy or hormonal therapy for prostate cancer, or could not provide informed consent. Purposive sampling sought men at different stages of AS.

The topic guide covered dietary fibre supplementation and proposed study procedures, including diet diaries and biological samples. It was developed using a published framework [20] and pilot-tested with three men on AS recruited through the UCAN Urological Cancer Charity centre.

Interviews were audio-recorded and transcribed verbatim. The transcripts were analysed using deductive thematic analysis guided by the Theoretical Framework of Acceptability [21]. Coding was undertaken using NVivo 12 Pro qualitative data analysis software (QSR International Pty Ltd, Melbourne, Australia). Themes were refined through repeated review and research-team debriefing.

The qualitative study was approved the West Midlands-Coventry and Warwickshire Research Ethics Committee (Project ID 340457) and NHS Grampian Research and Development (24/PR/0681), registered in Clinicaltrials.gov (NCT07790003) .

## Results

### Overall active surveillance cohort

Between January 2012 and December 2022, 420 men were enrolled in the AS pathway at NHS Grampian. The median age at diagnosis was 65 years (IQR: 60-69). Men were more commonly White Scottish/English (90.7%), married (69.8%) and from less deprived socioeconomic backgrounds (40.0% in SIMD Quintile 5). Family history of prostate cancer was documented in 10.5% of men. At diagnosis, 95.2% had a PSA <10 ng/mL (mean 4.4±3.1 ng/mL), and 99.8% had Gleason 3+3 (Grade Group 1) disease. Ninety five percent were classified as Cambridge Prognostic Group 1.

The median follow-up from active surveillance enrolment to discontinuation or censoring was 3.1 years (interquartile range [IQR] 1.6-5.1). During follow-up 208 men (49.5%) discontinued active surveillance for various reasons (see below), while 212 men (50.5%) were still on AS at last follow-up. No deaths were documented as being caused by prostate cancer while on AS, although nine men died while on AS from unrelated causes: six from cardiovascular disease and three from metastatic spread from other malignancies (lung, gastrointestinal tract, lymphoma). One further man died after leaving AS and receiving radiotherapy for intermediate-risk disease (no cause of death was documented, although his last assessment showed no evidence of metastatic disease). By Kaplan-Meier analysis, the median time to AS discontinuation was 4.9 years (95% Cl 4.4-5.6). The estimated probability of remaining on AS was 95.4% (95%CI: 92.9–97.1) at 1 year, 77.9% (95%CI: 73.5–81.7) at 2 years, 67.8% (95%CI: 62.8–72.3) at 3 years, 58.9% (95%CI: 53.5–63.9) at 4 years and 49.2% (95%CI: 43.4–54.7) at 5 years (**Figure 1**). Therefore, the percentage of men discontinuing AS after one year was 4.6% (95%CI: 2.9–7.1), after 2 years 22.1% (95%CI: 18.3–26.5); after three years, 32.2% (95% CI: 27.7–37.2) and after 5 years, 50.8% (95%CI: 45.3–56.6).

**Figure 1.**
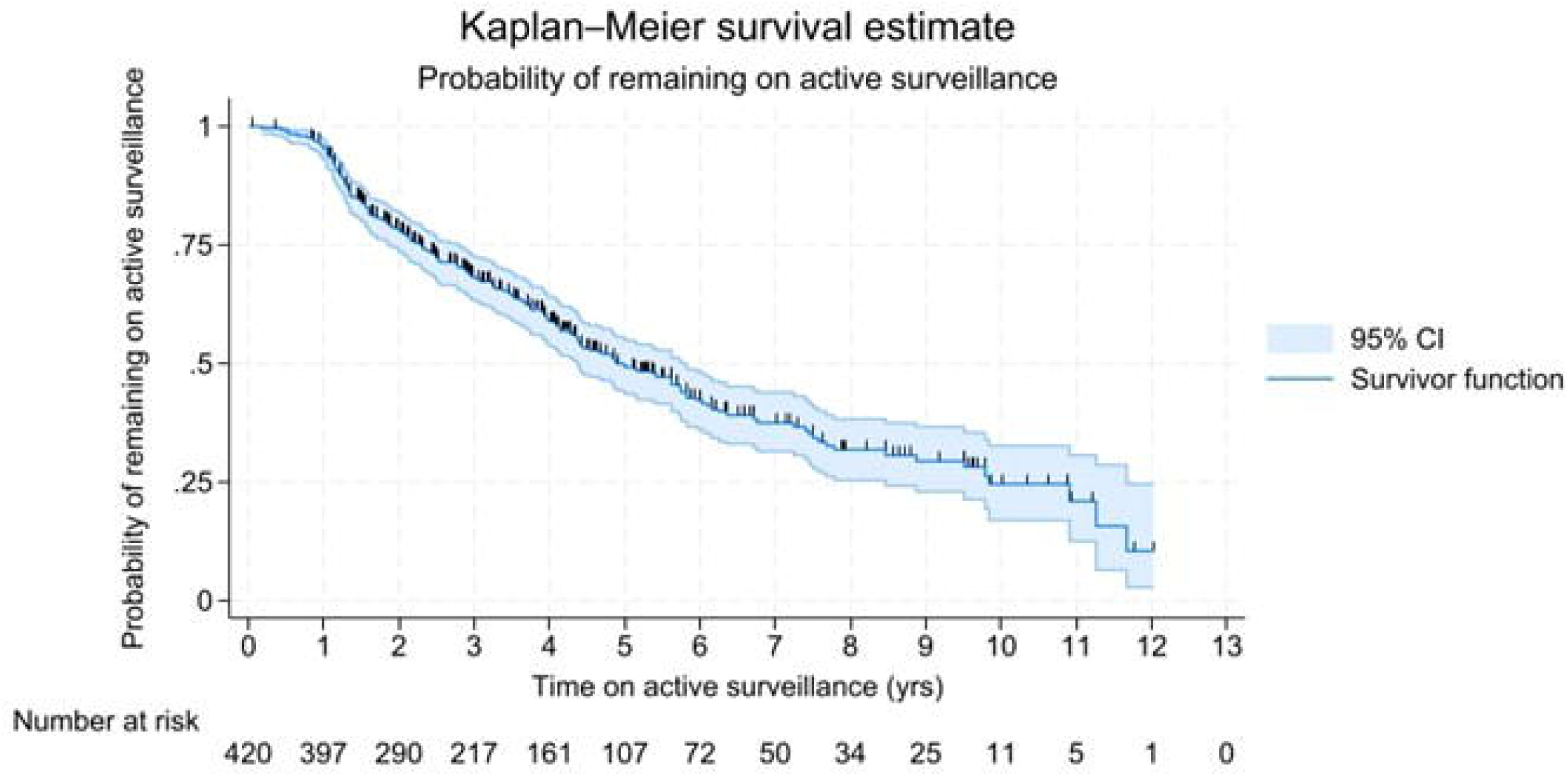
Kaplan-Meier estimate of the probability of remaining on active surveillance in the whole cohort. (N=420). The median time to discontinuation was 4.9 years (95% CI 4.4-5.6). Vertical marks indicate censoring; the shaded area shows the 95% CI. Numbers at risk are shown below the plot.

Disease progression accounted for 176 of 208 AS discontinuations (84.6%), occurring at a median of 2.1 years (IQR 1.3-3.8). Pathological progression was the most frequent mode of progression, accounting for 125 discontinuations (60.1%). Of these, 119 men upgraded to Grade Group ≥2, while six had increased pathological tumour burden without grade upgrading. Median time to pathological progression was 1.8 years (IQR 1.3-3.0).

Reasons for discontinuation unrelated to progression (**Table 1**) included patient choice (n=14, 6.7%), at median 3.9 years (IQR: 1.2-5.6), and transition to watchful waiting due to advancing age (n=18, 8.7%), at median 7.4 years (IQR: 3.9-9.5; median age 75.5 at time of transitioning; range of age 71-83).

**Table 1.** Reasons for active surveillance discontinuation (N=208).

| **Reason** | **n (%)** |
| --- | --- |
| Pathological progression | 125 (60.1) |
| Combined radiological and biochemical progression | 24 (11.5) |
| Radiological progression only | 7 (3.4) |
| Biochemical progression only | 20 (9.6) |
| Patient choice | 14 (6.7) |
| Transition to watchful waiting | 18 (8.7) |

### Comparisons between MRI-informed pathway and Biopsy-first pathway

Baseline demographic and clinical characteristics according to diagnostic pathway are summarised in **Table 2**. No statistically significant differences were seen between the MRI and biopsy-first pathways in age at diagnosis, PSA level, Gleason score distribution, Cambridge prognostic group classification, core positivity, or estimated tumour volume. The mean number of positive cores identified on diagnostic biopsy was higher in the MRI pathway than in the biopsy-first pathway (2.4 vs 1.9; p=0.04).

**Table 2.**
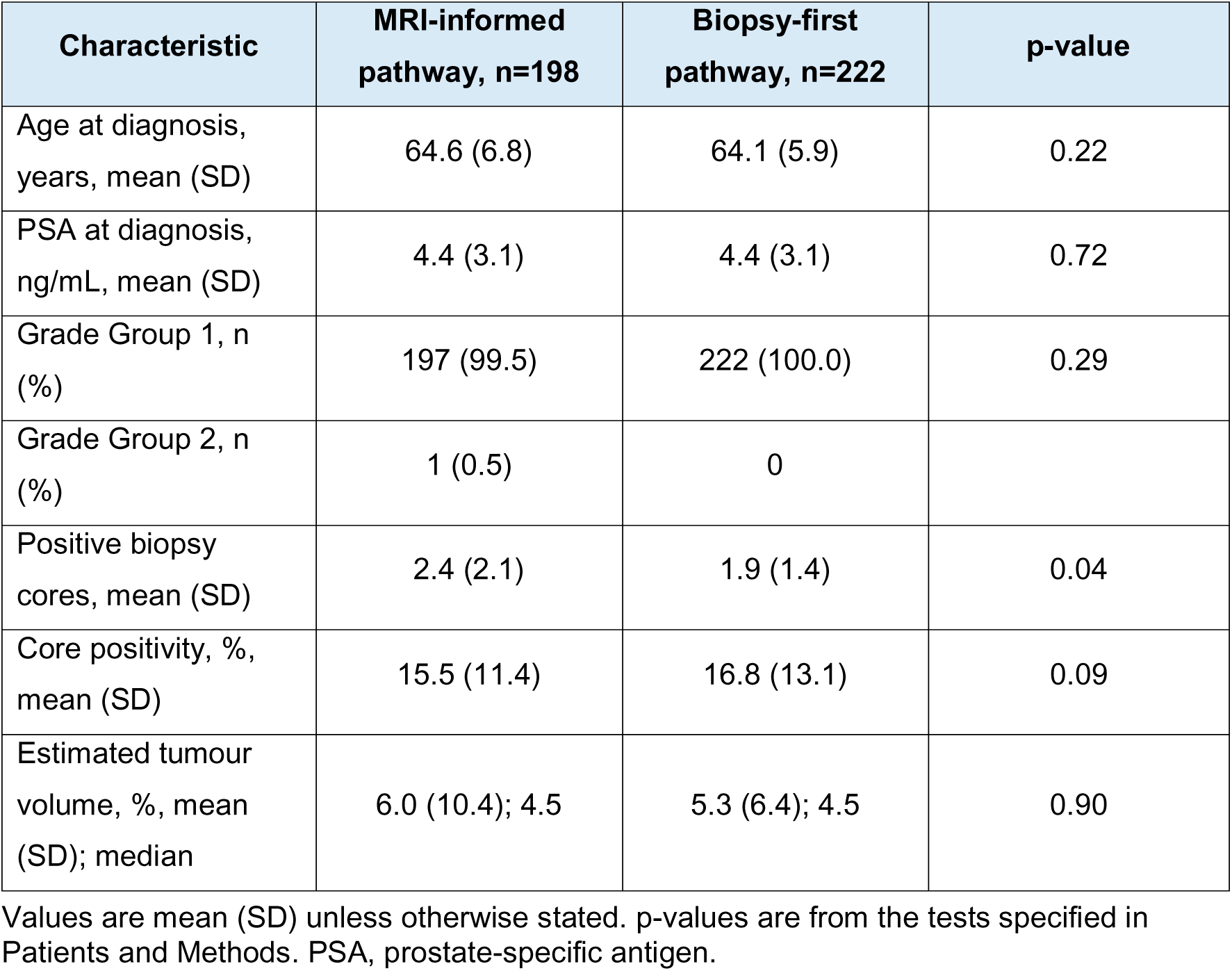
Baseline clinical characteristics by diagnostic pathway.

| Characteristic | MRI-informed pathway, n=198 | Biopsy-first pathway, n=222 | p-value |
| --- | --- | --- | --- |
| Age at diagnosis, years, mean (SD) | 64.6 (6.8) | 64.1 (5.9) | 0.22 |
| PSA at diagnosis, ng/mL, mean (SD) | 4.4 (3.1) | 4.4 (3.1) | 0.72 |
| Grade Group 1, n (%) | 197 (99.5) | 222 (100.0) | 0.29 |
| Grade Group 2, n (%) | 1 (0.5) | 0 |  |
| Positive biopsy cores, mean (SD) | 2.4 (2.1) | 1.9 (1.4) | 0.04 |
| Core positivity, %, mean (SD) | 15.5 (11.4) | 16.8 (13.1) | 0.09 |
| Estimated tumour volume, %, mean (SD); median | 6.0 (10.4); 4.5 | 5.3 (6.4); 4.5 | 0.90 |

Values are mean (SD) unless otherwise stated. p-values are from the tests specified in Patients and Methods. PSA, prostate-specific antigen.

The MRI-informed pathway included 198 men and the biopsy-first pathway 222 men **(Figure 2).** Median time to discontinuation was 4.1 years (95% CI 3.5–4.9) and 5.6 years (95% CI 4.9–6.5), respectively (log-rank p<0.01).

**Figure 2.**
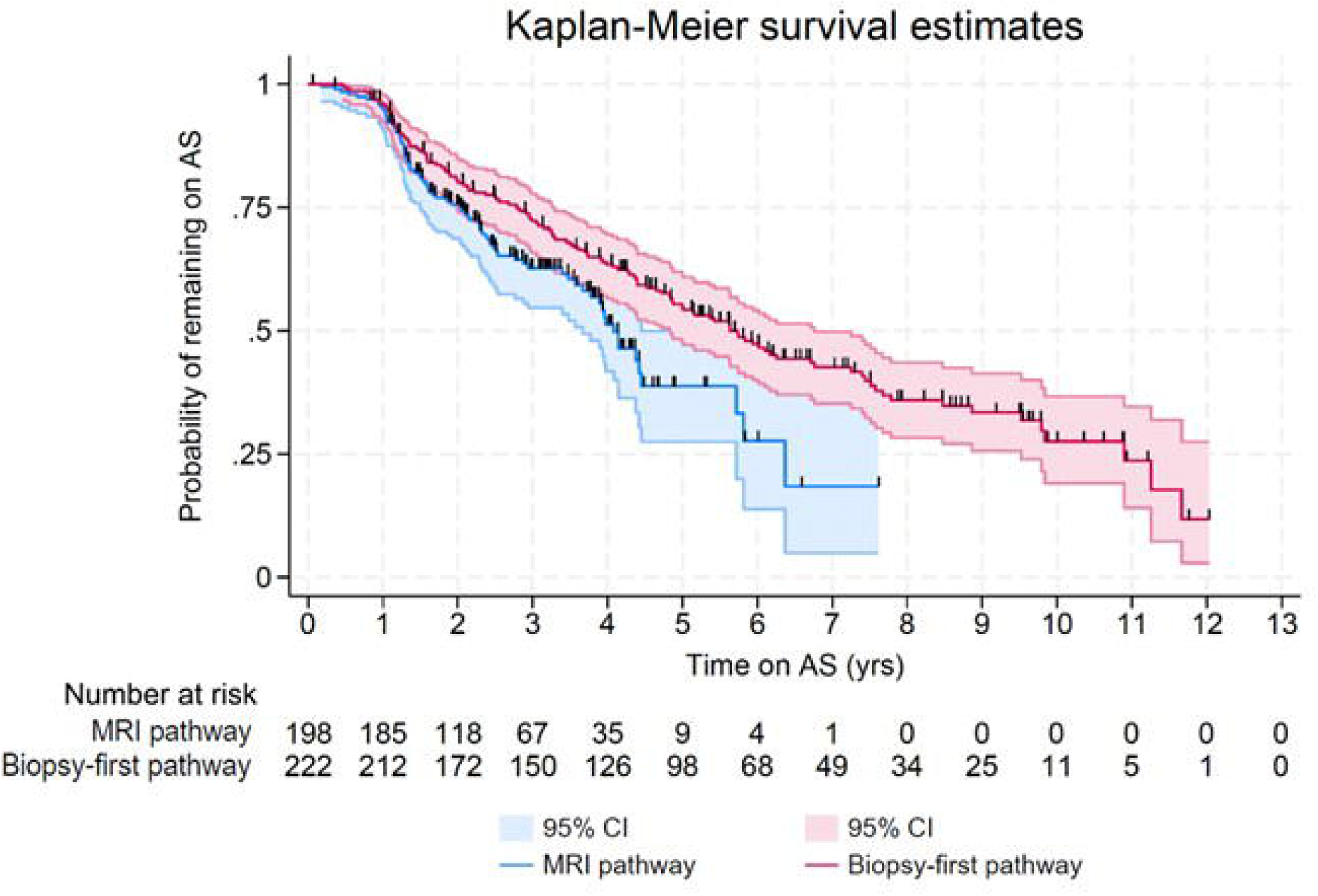
Kaplan-Meier estimates of the probability of remaining on active surveillance by diagnostic pathway. Vertical marks indicate censoring; shaded areas show 95% CIs.

Among men who discontinued AS, pathological progression accounted for 60 of 81 (74.1%) discontinuations in the MRI-informed pathway and 65 of 127 (51.2%) in the biopsy-first pathway (p=0.001). Pathological progression was identified earlier in the MRI-informed pathway (median 1.4 years, IQR 1.2-2.4) than in the biopsy-first pathway (2.1 years, IQR 1.4-3.9; p<0.01). In the MRI-informed pathway, 11 of 81 discontinuations (13.6%) involved radiological progression: 10 combined with biochemical progression and one due to radiological progression alone; corresponding numbers in the biopsy-first pathway were 14 and six, respectively (p=0.77 and p=0.25). Biochemical-only progression occurred in two and 18 men, respectively (p=0.01).

Transition to watchful waiting occurred in four of 198 men in the MRI-informed pathway and 14 of 222 men in the biopsy-first pathway. Because the MRI-informed pathway was introduced later and had shorter available follow-up, this difference was considered descriptive rather than a comparative pathway outcome.

Median time from diagnostic biopsy to first repeat biopsy did not differ between the MRI-informed and biopsy-first pathways (1.3 years [IQR 1.1-1.6] vs 1.3 years [IQR 1.1-2.0]; p=0.34). However, the interval between first and second repeat biopsies was shorter in the MRI pathway (median 2.0 years, IQR: 1.7-2.3) compared to the biopsy-first pathway (median 2.2 years, IQR: 2.0-2.9; p<0.01).

The MRI-informed pathway scheduled MRI at years 0, 1, 2 and 3 (MRI intervals extended to every two years thereafter), whereas MRI in the biopsy-first pathway was performed less systematically. Accordingly, the median interval from baseline to first repeat MRI was shorter in the MRI-informed pathway (1.2 years, IQR 1.0-1.4) than in the biopsy-first pathway (1.7 years, IQR 1.1-2.3; p<0.001).

Among 167 men in the MRI-informed pathway with recorded baseline PI/RADS/Likert scores, 44 had scores of 4-5 and 123 had scores of 2-3. Median time to AS discontinuation was shorter in men with scores of 4-5 than in those with scores of 2-3 (2.9 years (95% CI 2.3-3.5) vs 4.0 years (95% CI 3.7-4.3) (p<0.01; **Figure 3**)).

**Figure 3.**
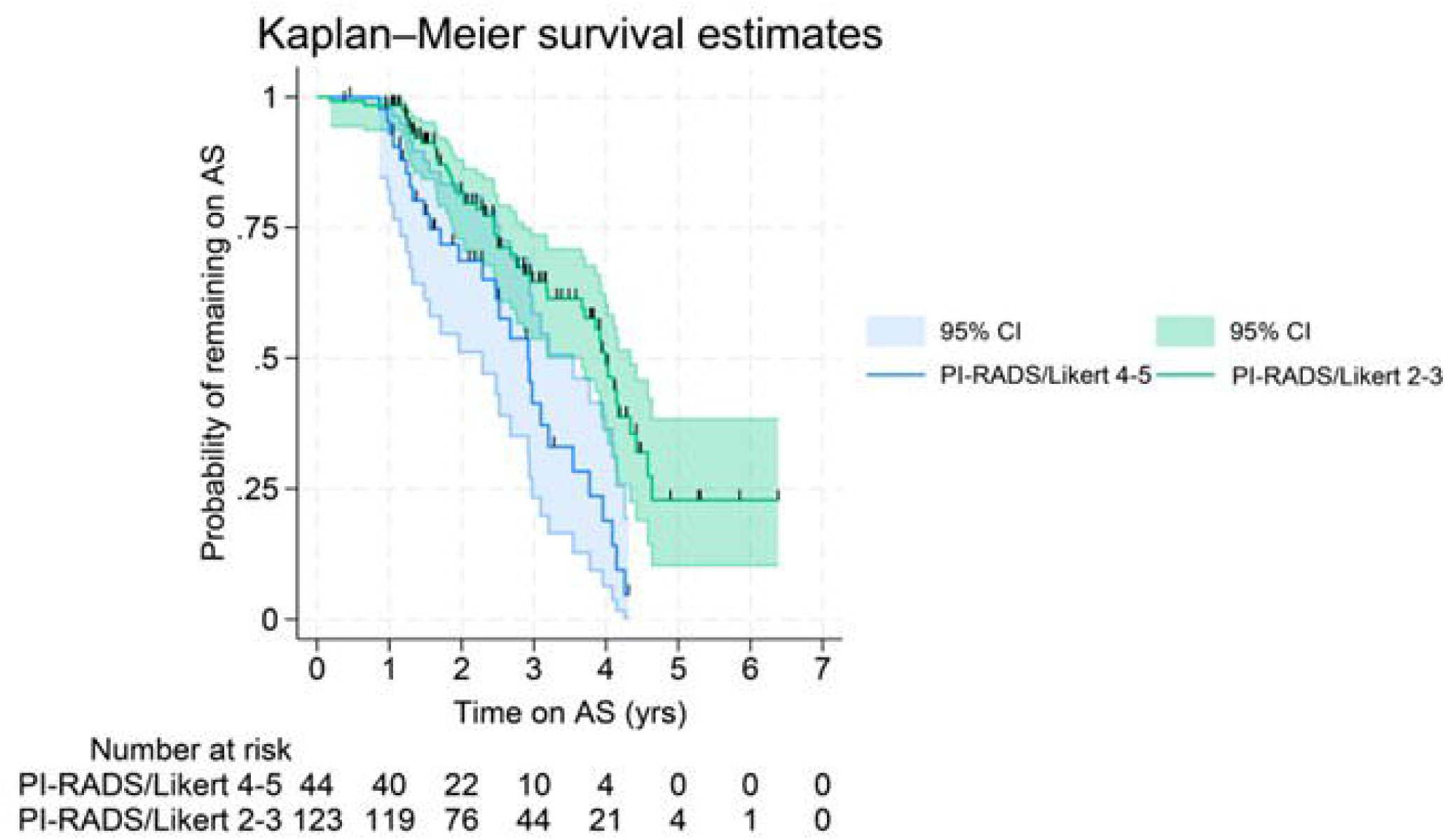
Kaplan-Meier estimates of the probability of remaining on active surveillance by baseline PI-RADS/Likert score. Vertical marks indicate censoring; shaded areas show 95% CIs. PI-RADS, Prostate Imaging Reporting and Data System.

Among 208 patients who discontinued AS, disease progression accounted for 176 (84.6%) discontinuations. Of 123 patients with PI-RADS/Likert 2–3, 30 (24.4%) discontinued active surveillance (AS), due to pathological progression 25 (83.3%), combined MRI and biochemical progression 2 (6.7%), and single cases of MRI-based progression alone (3.3%), patient choice and transition to watchful waiting. None discontinued for biochemical progression alone.

Of 44 patients with PI-RADS/Likert 4–5, 39 (88.6%) discontinued AS due to pathological progression 28 (71.8%), combined MRI and biochemical progression 5 (12.8%), patient choice 3 (7.7%), biochemical progression alone 2 (5.1%), and transition to watchful waiting 1 (2.6%). None discontinued because of MRI-based progression alone. Reasons for discontinuation did not differ significantly between PI-RADS 2-3 and 4-5 groups (p=0.607).

### Qualitative Study: Acceptability of Dietary Fibre Supplementation

Ten men on AS participated in semi-structured interviews (median age 71 years, range 55-76; median time since diagnosis 1.8 years, range 0.5-4.0). Thematic analysis using the Theoretical Framework of Acceptability revealed positive attitudes toward dietary fibre supplementation. Analysis of the semi-structured interviews yielded six themes corresponding to constructs of the TFA (affective attitude, burden, ethicality, intervention coherence, perceived effectiveness and self-efficacy). The seventh TFA construct, opportunity cost, did not prominently feature in participants’ responses.

Overall, participants expressed a predominantly positive affective attitude toward the proposed fibre supplement and demonstrated confidence in their ability to incorporate it, despite anticipating certain burdens associated with side effects and trial procedures. Many were enthusiastic about “giving it a go” and indicated they would be willing to try anything that might be beneficial for their health. As one man succinctly put it, *“Yeah, I mean if it helps then anything’s worth a… yeah” (Pt01),* a sentiment echoed by others who said they would be *“willing to try anything, really” (Pt02)* and were *“happy to give it a go” (Pt06).* Several noted that because they were already accustomed to regular doctors’ appointments and tests (through AS), adding a few extra tasks would be manageable. For instance, one participant shared that *“I go every three months to get a blood test for the PSA count… I’ve taken poo samples for bowel cancer… so I’ve done that in the past” (Pt02)*.

The burden theme captures the perceived effort and inconvenience associated with the intervention. While affective attitudes were largely positive, participants also identified various burdens (i.e. potential sources of effort, inconvenience or discomfort) that could arise from taking the supplement and participating in the trial.

*“If it was really, really bad I’d be reluctant to take it, but…I don’t think it would be too bad” (Pt05)*.

Importantly, most men viewed these burdens as manageable or conditional: mild difficulties were deemed acceptable in the pursuit of prostate cancer benefits, whereas more severe or sustained burdens might limit their willingness to continue.

*“Probably not [a problem], but it would depend once it started, how bad it was,”* one participant explained, adding with good humour, *“Just make sure the windows are open, I suppose” (Pt01)*.

Many men’s personal values aligned with the study aims, reflecting a sense of ethicality through altruism and duty to contribute. As one participant put it, *“If you’re going to be taking part in a study, you have to accept that you do anything you can to get the results… if you’re going to participate you might as well participate [fully].” (Pt01).* Another similarly commented, *“I’ll certainly give it a try… if it’s going to help other people in life I’d be quite happy with that.” (Pt10)*.

At the same time, participants described their baseline understanding and needs regarding the intervention (intervention coherence), as well as their beliefs about its likely benefit (perceived effectiveness). Virtually all participants reported that they had never heard of using a dietary fibre supplement in the context of AS in prostate cancer before it was introduced to them in this study. A couple of men mentioned general awareness of dietary fibre in nutrition (e.g. *“I have porridge in the morning”* as a source of fibre (Pt02)), but none had experience with fibre supplement products.

*“No one’s actually told me anything about it… I’ve never heard of them,”* one participant said, adding that “*if I knew about them I would most probably take them” (Pt04)*.

There was a prevailing attitude of *“I’m happy to do it, I just need to know what to do”.* As one man summarised, the men were generally ready to help “provided they understood why and how” to carry out each task.

Finally, participants discussed their confidence and potential challenges in adhering to the regimen over time (self-efficacy). Generally, men conveyed high confidence that they could adhere to the protocol, often drawing on their past experiences and personal choices.

*“If you were doing the study… [it’d be] par for the course, wouldn’t it? Yeah, definitely,”*

They discussed practical strategies for fitting the supplement and tasks into their daily lives. However, some also reflected on how sustaining these activities over a long period might become challenging, identifying conditions that could erode their initial confidence.

*“If it was just a matter of two or three months, then fine. If it was a year’s study… that might be a little bit onerous” (Pt09)*.

## Discussion

The median time to AS discontinuation was 4.9 years. This is broadly comparable with established AS cohorts, including the Toronto cohort and PRIAS, although direct comparison is limited by differences in eligibility criteria, surveillance protocols and outcome definitions [2,7].

Disease progression accounted for 176 of 208 (84.6%) discontinuations, representing 41.9% of all men enrolled. Pathological progression accounted for 125 discontinuations (60.1%), with 119 men upgraded to Grade Group ≥2, while six had increased pathological tumour burden without grade upgrading. Median time from AS enrolment to pathological progression was 1.8 years, suggesting that pathological reclassification at early repeat biopsy could inform future intervention studies. In the MRI-informed pathway, 11 of 81 (13.6%) discontinuations involved radiological progression.

No prostate cancer-specific deaths were recorded during observed follow-up. The ProtecT trial similarly reported low prostate cancer-specific mortality after 15 years across active monitoring, surgery, and radiotherapy, although active monitoring in ProtecT was not equivalent to contemporary protocol-driven AS [1].

### MRI Pathway and AS Discontinuation

Men managed through the MRI pathway discontinued AS sooner than those on the biopsy-first pathway. Pre-biopsy mpMRI improves localisation of suspicious lesions and assists targeted biopsy and risk stratification [3, 22,23].

At baseline, men in the MRI-informed pathway had a higher number of positive cores and surveillance also differed between pathways. The shorter interval between first and second repeat biopsies in the MRI-informed pathway was expected from the surveillance protocols: the biopsy-first pathway scheduled repeat biopsies at years 1 and 4, whereas the MRI-informed pathway scheduled repeat biopsy at approximately year 1, 2 and 3. This greater early surveillance intensity provided more opportunities to detect pathological upgrading earlier. MRI-informed assessment may improve early pathological reclassification, with MRI-targeted plus systematic confirmatory biopsy detecting more Grade Group _≥_2 disease than systematic biopsy alone, and the ASIST study reporting fewer subsequent AS failures after MRI-informed confirmatory assessment [24,25]. Therefore, the earlier pathological progression observed in our MRI-informed pathway may partly reflect improved detection and earlier reclassification of previously occult higher-grade disease rather than a greater underlying rate of biological progression.

Biochemical-only progression was less frequent in the MRI-informed pathway (2 vs 18 men, respectively), consistent with MRI-led AS studies in which PSA changes are interpreted alongside imaging and used to trigger further assessment rather than acting as an isolated marker of progression. Gallagher et al. used serial MRI and PSA dynamics to guide selective re-biopsy, while the prospective MRIAS study used MRI changes and PSA density to trigger surveillance biopsy [26, 27].

Exploratory PI-RADS/Likert analysis also identified shorter times remaining on AS for men with PI-RADS/Likert scores 4-5 compared with those with scores 2-3. The PI-RADS/Likert 4-5 subgroup represented approximately 44 of 167 with recorded scores, or roughly one quarter of men, providing a potentially feasible strategy for selecting participants for future dietary intervention studies, namely those most likely to discontinue AS earlier, with the caveat that the endpoint was discontinuation of AS for any reason, rather than disease progression alone.

Transitions to watchful waiting should also be interpreted cautiously. Watchful waiting reflects reduced suitability for curative treatment owing to age, comorbidity, or estimated life expectancy, rather than cancer progression. The biopsy-first cohort had longer potential follow-up than the more recently introduced MRI pathway cohort and therefore greater opportunity for men to age into eligibility for watchful waiting.

### Dietary Fibre Acceptability

The acceptability of dietary fibre interventions has been evaluated in other clinical populations, predominantly through measures of adherence and gastrointestinal tolerability rather than qualitative frameworks. In individuals at increased colorectal cancer risk, a 12-week rice bran and navy bean intervention achieved high retention and compliance, while a shorter intervention in colorectal cancer survivors reported no major gastrointestinal problems [28,29]. Higher-fibre dietary interventions have also been tolerated during pelvic radiotherapy [30]. These findings are consistent with our participants’ general willingness to consider dietary fibre supplementation but also highlight the importance of intervention burden. In our study, concerns extended beyond gastrointestinal effects to prolonged diet recording and repeated biological sampling. Future feasibility studies should therefore evaluate acceptability of the supplement and the accompanying study procedures separately.

Existing clinical evidence for dietary fibre interventions in prostate cancer is preliminary and generated in heterogeneous patient populations, with different stages of prostate cancer and different interventions. Only the Thomas et al study [15] was conducted in men AS; however, longer follow-up is required to determine whether observed differences in PSA progression translate into clinically meaningful differences in pathological progression or treatment.

The present qualitative findings support further feasibility work. Our participants highlighted that future study design should minimise practical burden, avoid overly intensive long-term diary requirements, coordinate sample collection with routine clinical follow-up where possible and use an intervention format acceptable to participants.

### Strengths and Limitations

Strengths of this study include a large regional cohort, detailed classification of reasons for AS discontinuation, direct comparison of historical surveillance pathways and integration of clinical outcomes with patient interviews. The combined inductive–deductive analysis strengthened confidence in the qualitative findings. Mapping the data to the Theoretical Framework of Acceptability (TFA) grounded the interpretation in an established theory, while inductive coding ensured that important themes outside the framework were not missed.

This study has several limitations. First, it was retrospective and single centre. Second, the pathway groups were treated during different periods and differed in diagnostic approach, surveillance schedules and available follow-up, limiting attribution of the observed differences specifically to pre-biopsy MRI. Diagnostic practice within the MRI-informed pathway was also heterogeneous, as MRI-targeted biopsy was not performed consistently in men with suspicious MRI findings. Third, AS discontinuation included several clinically distinct reasons for leaving surveillance. Finally, the PI-RADS/Likert analysis was exploratory and limited to men with recorded baseline scores.

### Clinical Implications and Future Research

These data provide a contemporary description of AS outcomes within a single university teaching hospital in the north east of Scotland and demonstrate the importance of distinguishing between pathological progression, biochemical or radiological triggers, patient choice and transition to watchful waiting when reporting discontinuation from surveillance. The comparison of diagnostic pathways suggests that the implementation of MRI-informed surveillance was associated with earlier AS discontinuation, influenced by changes in monitoring intensity and opportunities for pathological reclassification.

Men with a modest PSA elevation or low PSA density and a non-suspicious MRI (PI-RADS/Likert 1–2) may not undergo biopsy when overall clinical suspicion is low [3], whereas men with suspicious imaging or persistent clinical concern are more likely to be biopsied. As this audit included only men with biopsy-confirmed prostate cancer who subsequently entered AS, it did not capture men in whom biopsy was omitted. The MRI-informed cohort may therefore have been enriched for men with greater baseline suspicion and a higher likelihood of subsequent pathological reclassification. Earlier AS discontinuation may consequently reflect pre-enrolment selection, as well as differences in surveillance intensity and opportunities for detecting previously occult higher-grade disease.

For future dietary fibre research, the present findings support the development of suitable feasibility studies. Such studies should prioritise recruitment, acceptability, adherence, gastrointestinal tolerability, completion of study procedures and retention. Exploratory clinical or biological outcomes could include PSA kinetics, gut microbiota characterisation and patient-reported outcomes.

## Conclusions

Approximately half of men discontinued AS by 5 years, most often after pathological progression was detected. Earlier discontinuation in the MRI-informed pathway may reflect more intensive monitoring and changes in diagnostic practice as well as MRI use. Exploratory analysis of baseline PI-RADS/Likert 4-5 identified a sizeable subgroup with earlier all-cause AS discontinuation that may inform future recruitment into clinical trials of dietary fibre supplementation. Dietary fibre supplementation was acceptable in principle to men on AS.

## Data Availability

Anonymised data produced in the present study may be available upon reasonable request to the authors.

## Acknowledgements

This study formed part of DS’s MSc by Research project and was funded by Friends of ANCHOR. AEK holds the Friends of ANCHOR Clinical Chair in Oncology at the University of Aberdeen. We thank Kevin Wardlaw for his help in identifying the patient records to include in this study, and the nursing staff at the UCAN Centre and the staff of the Department of Urology at Aberdeen Royal Infirmary for their support.

