## Supplemental Material for "MRI-informed active surveillance outcomes and dietary fibre acceptability: the Aberdeen experience"

**Supplementary Material**

**Supplementary Table S1. Variables collected in the retrospective audit.**

| **Domain** | **Variables** |
| --- | --- |
| Demographics | Age; marital status; ethnicity; Scottish Index of Multiple Deprivation quintile; family history of prostate cancer. |
| Biochemical | PSA at diagnosis and during surveillance; PSA density; PSA doubling time. |
| Histopathology | Biopsy type; Gleason score/Grade Group; number of positive cores; percentage tumour involvement; estimated tumour volume. |
| Radiology | Prostate volume; PI-RADS/Likert score; lesion characteristics; PRECISE score on serial MRI. |
| Outcomes | AS enrolment and follow-up dates; AS status; date and reason for discontinuation; transition to watchful waiting; death and recorded cause. |

**Supplementary Table S2. Summary of the historical biopsy-first and more recent MRI-informed active surveillance protocols.**

| **Component** | **Biopsy-first protocol** | **MRI-informed protocol** |
| --- | --- | --- |
| Diagnostic pathway | Systematic biopsy without routine pre-biopsy MRI. | Pre-biopsy mpMRI whenever possible; targeted and/or systematic biopsy according to clinical practice. |
| PSA monitoring | Every 3 months for 2 years; every 6 months to 5 years; annually thereafter. | Every 3 months initially; every 6 months when stable. |
| Clinical review | Digital rectal examination followed the same schedule as PSA testing. | Clinical review every 6 months; digital rectal examination was later removed from routine surveillance. |
| MRI | Used selectively for staging or suspected progression. | MRI at 12, 24 and 36 months, then every 2 years. |
| Protocol biopsy | Years 1, 4, 7 and 10, then every 5 years. | Years 1, 2 and 3, then every 2 years. |
| Biochemical trigger | PSA doubling time <3 years could prompt discontinuation; 3-10 years prompted repeat biopsy unless recently performed. | PSA density and PSA kinetics considered alongside MRI and biopsy findings. |
